# Comparison of two commercial platforms and a laboratory developed test for detection of SARS-CoV-2 RNA

**DOI:** 10.1101/2020.07.03.20144758

**Authors:** Laura Mannonen, Hannimari Kallio-Kokko, Raisa Loginov, Anu Jääskeläinen, Pia Jokela, Jenni Antikainen, Paula Väre, Eliisa Kekäläinen, Satu Kurkela, Hanna Jarva, Maija Lappalainen

**Affiliations:** HUS Diagnostic Center, HUSLAB, Clinical Microbiology, University of Helsinki and Helsinki University Hospital, Finland; Translational Immunology Research Program and Department of Bacteriology and Immunology, University of Helsinki, Helsinki, Finland

## Abstract

Mitigation of the ongoing COVID-19 pandemic requires reliable and accessible laboratory diagnostic services. We evaluated the performance of one LDT and two commercial tests, cobas® SARS-CoV-2 (Roche) and Amplidiag® COVID-19 (Mobidiag), for the detection of SARS-CoV-2 RNA in respiratory specimens. 183 specimens collected from suspected COVID-19 patients were studied with all three methods to compare their performance. In relation to the reference standard, which was established as the result obtained by two of the three studied methods, the positive percent agreement (PPA) was highest for cobas® test (100%), followed by Amplidiag® test and the LDT (98.9%). The negative percent agreement (NPA) was lowest for cobas® test (89.4%), followed by Amplidiag® test (98.8%) and the highest value was obtained for LDT (100%). The dilution series conducted for specimens, however, suggests significantly higher sensitivity for the cobas® assay in comparison with the other two assays and the low NPA value may be due to the same reason. In general, all tested assays performed adequately. Both the time from sample to result and hands-on time per sample were shortest for cobas® test. Clinical laboratories need to be prepared for uninterrupted high-throughput testing during the coming months in mitigation of the pandemic. To secure that, it is of critical importance for clinical laboratories to maintain several simultaneous platforms in their SARS-CoV-2 nucleic acid testing.

## Introduction

Mitigation of the ongoing COVID-19 pandemic requires reliable and accessible laboratory diagnostic services. The specific diagnosis of SARS-CoV-2 infection relies on molecular methods, especially on RT-PCR, although other technologies including serologic immunoassays are emerging^1,2^. The first methods for SARS-CoV-2 detection were laboratory developed RT-PCR tests (LDTs), one of the first methods published was described by Corman et al.^3^. This method was later endorsed by WHO and widely implemented in clinical laboratories. Roche Molecular Systems (Branchburg, NJ, USA) cobas® SARS-Cov-2 test was the first commercial test to get EUA from FDA on March 12 2020. Since then (as of June 25, 2020) more than 100 commercial molecular IVD-tests have been granted the FDA or other national authorities’ EUA and/or CE-mark^4^. Both LDTs and commercial tests have been set up in a high time pressure. Therefore, it is of great importance to evaluate the tests in clinical laboratory settings. In this study we evaluated the performance of one LDT and two commercial tests, namely cobas® SARS-CoV-2 (Roche) and recently CE/IVD marked Amplidiag® COVID-19 (Mobidiag, Espoo, Finland), for the detection of SARS-CoV-2 RNA in respiratory specimens.

## Material and Methods

### Specimens

Altogether 237 respiratory tract specimens referred to Helsinki University Hospital Laboratory (HUS Diagnostic Center, HUSLAB), Department of Virology and Immunology, Finland were included in this study. 54 specimens collected from patients with respiratory symptoms in 2018-2020 were used to verify analytical specificity of the tests. The specimens from 2020 were negative for SARS-CoV-2 RNA by the LDT. 183 specimens collected from suspected COVID-19 patients in 2020 were studied with all three methods to compare the performance of the tests in detecting SARS-CoV-2 RNA. Although most of the specimens tested were nasopharyngeal swabs, also oropharyngeal and nasal swabs were included due to the global shortage of the swab sticks needed for nasopharyngeal sampling. The specimens were collected either in Copan UTM (Copan, Brescia, Italy) or in tubes containing 0.9% saline due to the global shortage of Copan UTM tubes. The suitability of the 0.9% saline tubes as alternative to viral transport media for SARS-CoV-2 testing has been shown before^5^.

The specimens (n=183) deployed to compare the performance of the three studied RT-PCR methods comprised of two sets of specimens collected within two time frames. The first set of specimens (n=37) was part of the material used for the initial verification of cobas® SARS-CoV-2/Amplidiag® COVID-19 tests. These specimens were collected between March 5 and March 18, 2020, and the SARS-CoV-2 positive specimens (n=18) represent virus strains from the early epidemic. The second set of specimens (n=146) were collected between May 4 and May 8 2020, and the positive specimens (n=90) thus represent strains from a declining phase of the epidemic in Finland. Specimens were stored at -20 °C/-70 °C after initial analysis and were thawn upon analysis. Specimens were not thawn more than twice before RT-PCR.

In addition, QCMD 2020 Coronavirus Outbreak Preparedness EQA Pilot Study proficiency samples (Glasgow, Scotland, UK) were used to evaluate the performance of the methods.

### Inactivation and lysing of the specimens

All specimens were inactivated in a biosafety cabinet in a biosafety level 2 laboratory that has a negative pressure. At the beginning of the epidemic FFP3 mask, protective glasses and protective clothing were worn during working in the laboratory. Later in the epidemic visor and surgical mask replaced the FFP3 masks and protective glasses.

#### Laboratory developed test (LDT), HUSLAB

The possible SARS-CoV-2 in the specimen was inactivated by adding 250 µl of MagNA Pure Lysis/Binding Buffer (Roche Diagnostics GmbH, Mannheim, Germany) to 250 µl of patient specimen. Lysates were incubated for a minimum of 10 minutes before processed further.

#### Cobas® SARS-CoV-2

If needed, the specimens were first equilibrated to room temperature, after which the possible SARS-CoV-2 in the specimen was inactivated by adding 350 µl of MagNA Pure Lysis/Binding Buffer (Roche Diagnostics GmbH, Mannheim, Germany) to 350 µl of patient specimen. Lysates were incubated for a minimum of 10 minutes before processed further.

#### Amplidiag® COVID-19

The possible SARS-CoV-2 in the specimen was inactivated either by adding 600 µl of the specimen to an eNAT-tube (Copan) or 360 µl to an mNAT-tube (Mobidiag). eNAT tubes were incubated for a minimum of 30 minutes and mNAT-tubes for a minimum of 5 minutes before processed further.

### Molecular methods evaluated in the study

#### Laboratory developed test (LDT), HUSLAB

The real-time LDT SARS-CoV-2 RT-PCR used in this study is a modification of the method published by Corman et al.^3^. The test is suitable for detection of SARS-CoV-2 RNA from sputum, nasopharyngeal/tracheal aspirates, nasal, nasopharyngeal and oropharyngeal swab specimens and faeces. Initially, all target genes (E, RdRP and N) were included in the diagnostic assay. In addition, a PCR for beta-globin gene^6^ was performed in order to verify successful sampling, extraction and PCR. Full-length SARS-CoV *in vitro* transcript was used as a positive control. When the epidemic spread and there was suddenly a high demand of testing in combination with the global shortage of supplies, we first dropped out the E-gene-PCR, because of the occasional unspecific positive signal obtained from negative controls/specimens^7^. Later also RdRP was excluded and the diagnostics were continued with the N-gene-PCR only. The N-gene-PCR was chosen because a dilution series indicated better sensitivity for the N-gene-PCR over RdRP-PCR, although earlier ct-values were gained for RdRP-PCR. Also, findings from low positive specimens suggested better sensitivity for N-gene-RT-PCR (data not shown).

Nucleic acids were extracted from 450 µl respiratory specimen lysate using the MagNA Pure Viral NA SV 2.0 Kit (Roche Diagnostics GmbH, Mannheim, Germany), with the MagNA Pure 96-instrument, and eluted in 50 µl of the elution buffer.

Real time RT-PCR was performed using the SuperScript III Platinum One-Step qRT-PCR Kit with 600 nM of the forward primer CACATTGGCACCCGCAATC, 800 nM of the reverse primer GAGGAACGAGAAGAGGCTTG and 200 nM of the probe FAM-ACTTCCTCAAGGAACAACATTGCCA-BBQ^3^. RT-PCR reaction was performed on Stratagene Mx3005p PCR instrument (Agilent Technologies Inc, Santa Clara, CA, United States). Positive and negative controls were included in each run. Five µl of the extracted eluate was subjected to 25 µl PCR-reaction with cycling conditions of: 1 cycle of 55 °C for 20 min, 1 cycle of 95 °C for 15 min, followed by 45 cycles of 94 °C for 15 sec and 58 °C for 40 sec.

#### Cobas SARS-CoV-2 test

The cobas® SARS-CoV-2 test (Roche Molecular Systems) is a qualitative test for fully automated cobas® 6800/8800 platforms. The test is validated to detect SARS-CoV-2 RNA from nasal, nasopharyngeal and oropharyngeal swab specimens. The test amplifies two SARS-CoV-2 targets: orf1ab (Target 1), which is specific for SARS-CoV-2 and a conserved region of the E-gene (Target 2), which is pan-Sarbeco specific and detects also SARS-CoV and other Sarbecoviruses currently unknown to infect humans. In addition, the test includes an internal RNA-control (IC), which is added to the specimens before extraction. The test includes also positive and negative controls, which are processed the same way as the samples.

600 µl of specimen lysate was subjected to cobas® 6800 system. Testing was performed according to the manufacturer’s instructions apart from the inactivation step.

#### Amplidiag COVID-19 test

The Amplidiag® COVID-19 test (Mobidiag) is a qualitative test for the detection of SARS-CoV-2 RNA in nasopharyngeal specimens. The test amplifies two SARS-CoV-2 targets: orf1ab and N-gene, which are both specific for SARS-CoV-2. In addition, the test amplifies the human RNase P gene (RP), which serves as a sampling control. The test includes positive and negative control.

eNAT or mNAT tubes were subjected to Amplidiag® Easy system, which extracts nucleic acids and does the PCR setup. The PCR plate was then transferred to a real time PCR-machine (CFX-96, Bio-Rad, Hercules, CA, USA), which contains the Amplidiag® Analyzer software. The Amplidiag® Analyzer software transfers directly data from Amplidiag® Easy system and automates the result interpretation. Testing was performed according to the manufacturer’s instructions, apart from the specimen type (validated only for nasopharyngeal specimens) and inactivation step with eNAT-tubes.

A new version of the Amplidiag Easy Script Package (v. 5.1.0) and Amplidiag COVID-19 Kit Configuration (v2-0-1) were introduced by the manufacturer during the evaluation. The new version is not collecting data from the first 10 amplification cycles and therefore the obtained ct values are 10 cycles less than with the old version. The new software version was introduced in order to get rid of the drift problems seen with the orf1ab target (false positive interpretations of the old software). Since the new software did not allow analysis of the old data, 147 specimens from this evaluation were rerun from the same mNAT tubes within 5 days from the first run.

### Workflow evaluation of the studied tests

We compared the turnaround time (TAT), hands-on time and capacity of the tests. An experienced lab technician performed all three tests and measured time needed for each stage by stopwatch. The specimen inactivation step was excluded from this analysis.

Verification of the performance of all three studied methods on specimens collected to Copan UTM versus 0.9 % saline and using eNAT versus mNAT in the lysing/inactivation step with Amplidiag COVID-19 test

When the epidemic started, the specimens were collected to Copan UTM tubes (3 ml), but later to tubes containing 1.5 ml of 0.9% saline, because of the global shortage in supply of the Copan tubes. In order to verify that the used methods were performing at the same level two independent dilution series were constructed: (i). A patient specimen known to contain

SARS-CoV-2 RNA originally collected to Copan tube was diluted to pooled Copan-collected negative specimen. (ii). A patient specimen known to contain SARS-CoV-2 RNA originally collected to 0.9% saline was diluted to pooled 0.9% saline-collected negative specimen. The ct-value for SARS-CoV-2 detection by RT-PCR was around 24 for both the Copan-collected and saline-collected original positive specimens. A dilution series of 1:10 - 1:100000 for both specimens (in Copan UTM or saline) in respective specimen pools were constructed and analyzed with all three studied methods.

Due to the shortage of the eNAT tubes a change to mNAT tubes was necessary. This change was evaluated with a small number of specimens. Eleven specimens defined positive by the cobas® test were inactivated/lysed in parallel by pipetting the specimen to an mNAT and eNAT tube and analyzed in the same PCR run.

### Analytical specificity

The analytical specificity of the tests was studied by analyzing patient specimens containing other respiratory viruses. Altogether 54 specimens were included, but not all samples were run with all three methods. Some of the specimens contained several viruses (n=11). In four specimens, the presence of a respiratory virus nucleic acid was defined by xTAG RVP Fast (Luminex Diagnostics, Toronto, Canada) assay, and in 50 specimens by the Allplex Respiratory Panel 1/2/3 (Seegene, Seoul, Republic of Korea). The specimens were chosen to contain moderate to high concentrations of viral nucleic acids (ct-value <30 for specimens by Seegene test or high mean fluorescence intensity for Luminex test). The specimens were collected during 2018-2020 representing recent virus strains in Finland. The specimens are described in Table 1.

**Table 1.** 54 samples positive for other respiratory viruses tested with the assays evaluated.

| Clinical specimens with known viruses | Number of analyzed specimens |  |  |
| --- | --- | --- | --- |
|  | cobas® SARS-CoV-2 test | COVID-19 test | LDT |
| HCoV-229E | 2 | 2 | 2 |
| HCoV-OC43 | 3 | 3 | 3 |
| HCoV-NL63 | 4 | 6 | 4 |
| Influenza A | 2 | 2 | 4 |
| Influenza B | 2 | 2 | 4 |
| Rhinovirus | 3 | 3 | 5 |
| Enterovirus | 2 | 3 | 2 |
| Respiratory syncytial virus | 3 | 1 | 2 |
| Parainfluenza 1 virus | 2 | 3 | 2 |
| Parainfluenza 2 virus | 2 | 1 | 1 |
| Parainfluenza 3 | 2 | 2 | 2 |
| Parainfluenza 4 | 2 | 2 | 2 |
| Human metapneumovirus | 2 | 2 | 1 |
| Adenovirus | 2 | 1 | 2 |
| Human bocavirus | 2 | 3 | 3 |
| Total clinical specimens | 35 | 36 | 39 |

### Statistical methods

The positive percent agreement (PPA) and negative percent agreement (NPA) including two-sided 95% confidence intervals (CI) were calculated with an on-line MEDCALC® tool (https://www.medcalc.org/calc/diagnostic_test.php). The overall agreement of the evaluated assays was evaluated by the kappa value, which was calculated (including two-sided 95% CI) with an on-line QuickCalcs tool (https://www.graphpad.com/quickcalcs/kappa1/). Kappa values were interpreted as follows: No agreement (< 0), slight agreement (0-0.20), fair agreement (0.21-0.40), moderate agreement (0.41-0.60), substantial agreement (0.61-0.80), and almost perfect agreement (0.81-1). All values were calculated relative to the reference standard, which was established as the result obtained by two of the three studied methods. The significance of the difference of the ct values obtained by Amplidiag® test from eNAT vs mNAT tubes was estimated by two tailed students T-test. P-value < 0.05 was considered significant.

## Results

### Clinical performance of the evaluated assays

Altogether, 183 specimens were analyzed by all three evaluated methods. One specimen gave an invalid result by cobas® SARS-Cov-2 test (0.5%), two specimens had an incorrect amount of specimen lysate in LDT and were therefore considered invalid (1.1%), and 10 specimens gave failed results by Amplidiag® COVID-19 test (5.5%). The failed/invalid specimens were excluded from the agreement analysis. In relation to the reference standard the positive percent agreement (PPA) was highest for cobas® test (100%), followed by Amplidiag® test (98.9%) and the LDT (98.9%). Negative percent agreement (NPA) was lowest for cobas® test (89.4%), followed by Amplidiag® test (98.8%) and the best specificity value was obtained for LDT (100%). The overall agreement as defined by kappa-value was excellent for all studied tests (Table 2).

**Table 2.** The positive percent agreement (PPA), negative percent agreement (NPA) and concordance of the evaluated assays relative to the reference values (“consensus result” was used as a reference value and was defined as the result obtained by at least two of the three studied methods).

| Test |  | Reference value |  | kappa | PPA (95% CI) | NPA (95% CI) |
| --- | --- | --- | --- | --- | --- | --- |
|  |  | + | - |  |  |  |
| LDT | + | 87 | 0 | 0.988 (0.966-1.0) | 98.9% (93.83-99.97) | 100% (95.75-100) |
|  | - | 1 | 85 |  |  |  |
| Cobas | + | 88 | 9 | 0.896 (0.830-0.962) | 100% (95.89-100) | 89.4% (80.85-95.04) |
|  | - | 0 | 76 |  |  |  |
| Amplidiag + | + | 87 | 1 | 0.977 (0.945-1.0) | 98.9% (93.96-99.97) | 98.8% (93.62-99.97) |
|  | - | 1 | 84 |  |  |  |

### Workflow analysis of the tests

The TAT for the cobas® test was 3 h 30 min for 80 samples; for Amplidiag® test 3 h 30 min for 48 samples; and for the LDT test 4 h 30 min for 93 samples. The hands-on-time was shortest for the cobas® test (Table 3).

**Table 3.** Workflow evaluation

|  | cobas® SARS- | Amplidiag® COVID- |  |
| --- | --- | --- | --- |
|  | CoV-2 test | 19 test | LDT |
| Throughput (one run) | 93 (80*) | 48 | 93 |
| Throughput in 8 hr shift† | 372 | 96 | 186 |
| Hands-on time (one run) | 50 min | 1 hr 25 min | 2 hr 30 min |
| Turnaround time | 3 hr 15 min | 3 hr 30 min | 4 hr 30 min |

\*Due to laboratory process reasons only 80 specimens were run during the time measurements, even though the maximum capacity is 93 specimens + 3 controls
†Throughput in 8 hour shift was calculated with one operator and one set of instruments

### Performance comparison of the evaluated methods on specimens collected to Copan UTM versus 0.9 % saline

Two independent dilution series of two positive specimens were constructed to 0.9% saline and copan UTM in respiratory specimen matrix to verify that specimens collected to Copan and saline perform at the same level with the evaluated methods. The results indicate similar performance of all three methods independent of the collection media used (Table 4 and Figure 1). However, the data suggest higher overall sensitivity for the cobas® test as the dilution 1:20000 was still positive for both RT-PCR targets in the cobas® test, while 1:200-1:1000 was the last dilution yielding positive test results in the LDT and in the Amplidiag® test. The experiment was repeated with similar results. In the repeated experiment the last dilution with positive result with LDT and Amplidiag® test was 1:100 (both for samples in saline and Copan), while positive result was obtained for both targets in 1:10000 dilution by the cobas® test.

**Table 4.**
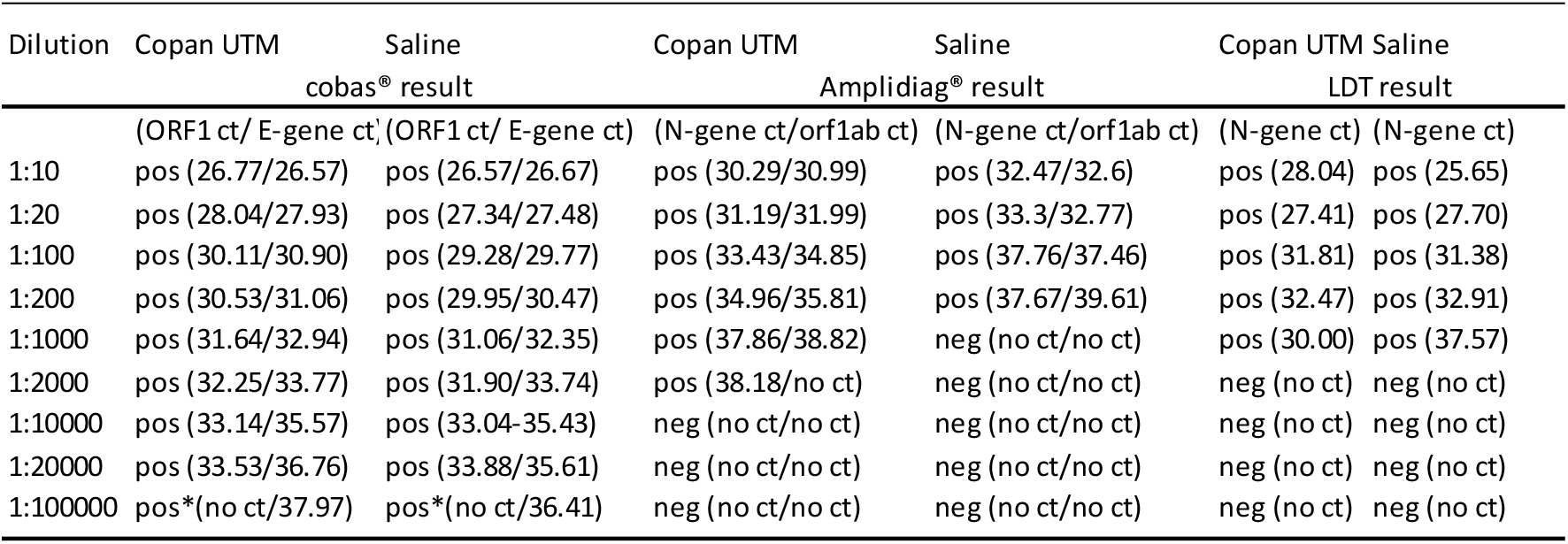
The comparison of results obtained from dilution series of SARS-CoV-2 in Copan UTM tubes and saline tubes.

**Figure 1.**
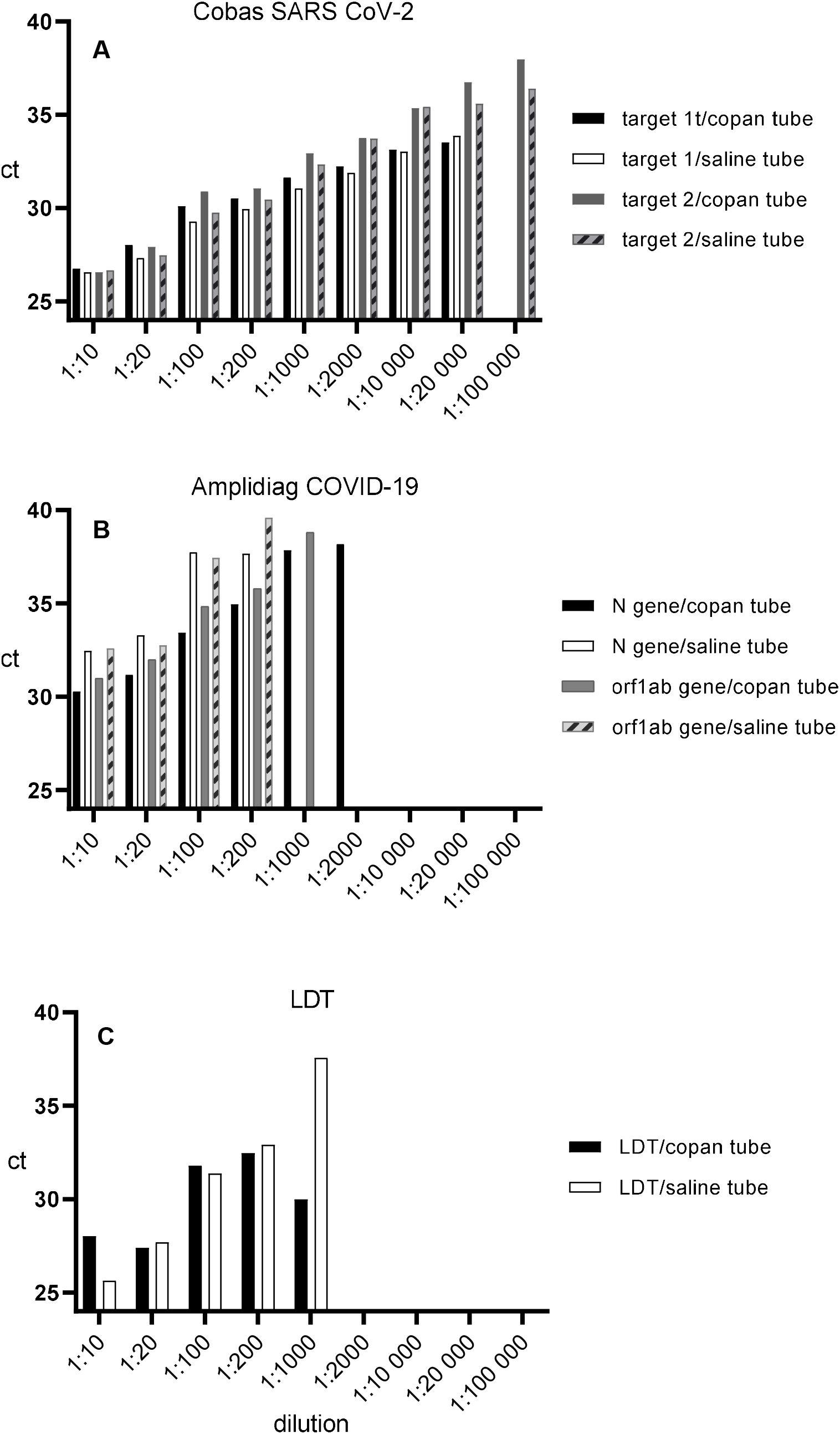
Comparison of the performance of the three studied methods using a dilution series in Copan UTM or 0.9% saline tubes. A known positive sample was diluted in either pooled Copan-collected negative specimen or saline-collected negative specimen. A) Cobas® SARS-CoV-2 test results. The test was positive for both targets at a dilution 1:20 000 for both Copan and saline samples. Target 2 was detected with both tubes still in dilution 1:100 000. B) Amplidiag® COVID-19 results. Amplidiag test was positive for both targets in Copan and saline tubes at 1:200 dilution but at 1:1000 and 1:2000 only in Copan tubes (positive for both targets at 1:1000 and only N-gene positive at 1:2000). No target was detected at dilutions 1:10 000 – 1:100 000. C) LDT assay. No target was detected at dilutions 1:2000-1:100 00. The Y axis is modified to show only ct’s 24-40 to allow for better visualization of the differences.

### Verification of eNAT versus mNAT tubes in the lysing/inactivation step with Amplidiag COVID-19 test

Of the 11 specimens defined positive by the cobas® test, 8 were positive with Amplidiag® test independent of the tube used for lysing/inactivation. Of the 8 specimens positive by the Amplidiag® test the difference in N gene and orf1ab target ct values were calculated and for the RNase P gene (sampling control) the difference was calculated from all 11 specimens. The mean difference in ct values was 0.77 for the N gene (P=0.004), 0.87 (P=0.189) for the orf1ab target and 1.98 for the RNase P-gene (P=0.026).

### Analysis of QCMD proficiency panel

QCMD 2020 Coronavirus Outbreak Preparedness EQA Pilot Study proficiency samples were used to evaluate the performance of the evaluated methods. The cobas® test and LDT were in 100% agreement with the QCMD expected results. Amplidiag® test failed to identify one SARS-CoV-2 positive result with SARS-CoV-2 concentration of 3.3 log_10_ copies/ml (Table 5). However, the test identified correctly another sample containing SARS-CoV-2 RNA 2.3 log_10_ copies/ml. Amplidiag® test reported four samples as failed because the EQA samples lack human RNase P target needed for valid negative results with the test.

**Table 5.**
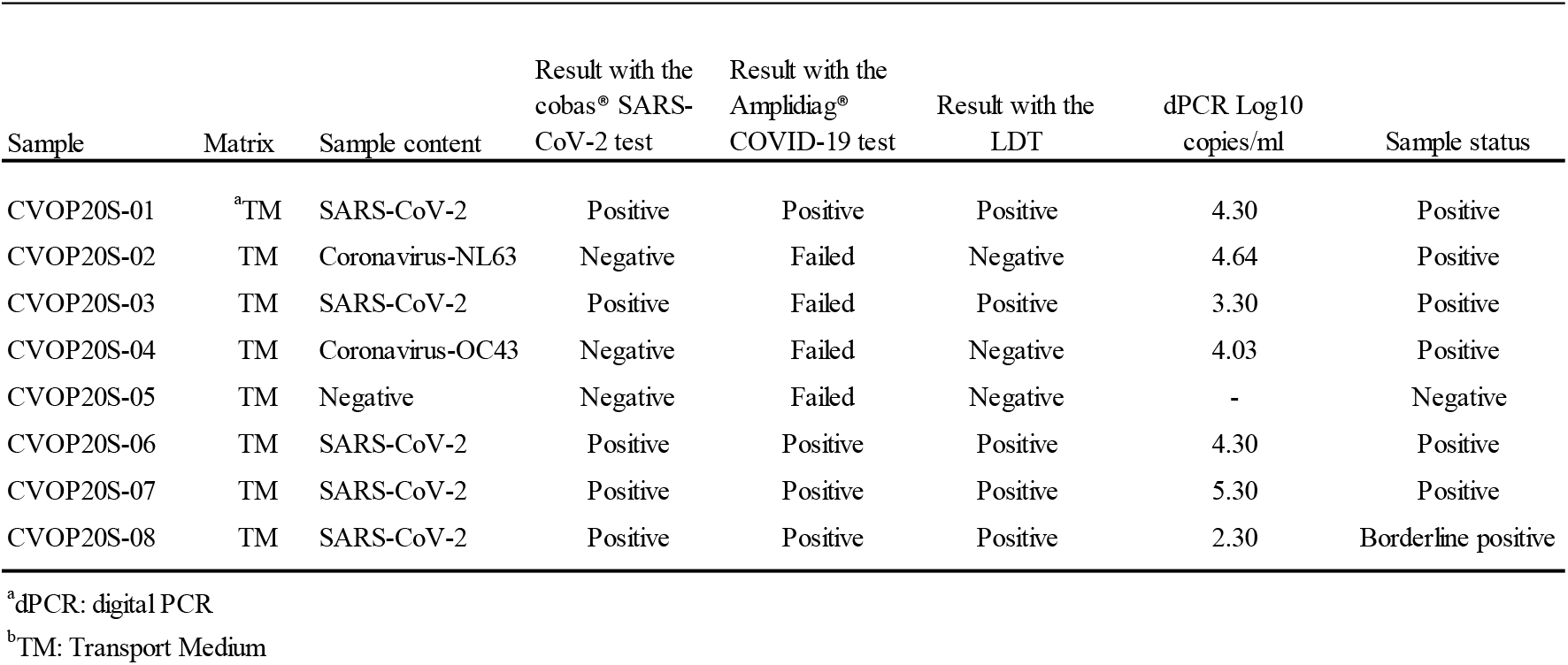
The results of QCMD 2020 Coronavirus Outbreak Preparedness EQA Pilot Study sample panel with the evaluated methods.

### Analytical specificity

None of the studied SARS-CoV-2 tests gave positive result from patient specimens containing other respiratory viruses (Table 1).

### Discrepant analyses

Altogether, there were 12 discrepant results. In one specimen, which only gave a positive result with Amplidiag® the positive signal was obtained for the orf1ab target with ct value of 40.8. However, in the amplification curve provided by Amplidiag® analyzer software no specific amplification could be seen. Instead of the expected logarithmic rise in the fluorescence signal, only a slow rise in background signal was seen (data not shown). For 1 specimen a positive result was obtained with cobas® and Amplidiag® tests, but a negative result with LDT. In one specimen positive results were obtained for cobas® test and LDT, while the Amplidiag® test gave a negative result. In that case the specimen had gone through 2 freeze-thaw cycles before it was analyzed by Amplidiag® test, but only one or none when analyzed by cobas® or LTD tests, respectively. In 9 specimens a positive result was obtained only by cobas® test. However, positive result was obtained for two specimens, when rerun with the new Amplidiag® COVID-19 software version. In all 9 specimens positive signal was obtained for both amplification targets of the test, but the ct values were high, >30 for 8/9 specimens. (Table 6A).

**Table 6.** Discrepant analysis: A. Discrepancies in the results between the evaluated methods. B. Discrepancies in the results between specimens analyzed with the old and the new version of Amplidiag software. Note that 10 ct cycles must be added to the ct-value obtained with the new version of the software in order to compare the values to the ct-values obtained by the old version.

**A**
| Number of specimens | Reference standard result | cobas® SARS-CoV-2 test result | LDT result | Amplidiag® COVID-19 test result | Comments |
| --- | --- | --- | --- | --- | --- |
|  |  | (ORF1 ct/ E-gene ct) | (N-gene ct) | (N-gene ct/orf1ab ct) |  |
| 1 | neg | neg (no ct) | neg (no ct) | pos (no ct/40.8) | Incorrect interpretation of the amplification curve by Amplidiag® Analyzer |
| 1 | pos | pos (33.7/34.9) | neg (no ct) | pos (no ct/38.1) |  |
| 1 | pos | pos (32.3/33.3) | pos (30.46) | neg (no ct) | The specimen had gone through two freezing-thawing cycles when analyzed by Amplidiag® and only one with cobas® test and non with LDT |
| 9 | neg | pos (28.6-34.4/29.1-37.9) | neg (no ct) | neg (no ct) | All specimens positive by cobas® only had a late ct value. Apart from one specimen, all had ct value > 30. Two of the specimens gave positive result when specimens were rerun with the new Amplidiag software. |

**B**
| Number of specimens | Amplidiag® result with old software | Amplidiag® result with new software | Comments |
| --- | --- | --- | --- |
|  | (N-gene ct/orf1ab ct) | (N-gene ct/orf1ab ct) |  |
| 1 | pos (no ct/40.8) | neg (no ct/no ct) | Incorrect interpretation of the amplification curve by old Amplidiag® software version |
| 4 | pos (no ct-38.1-no ct/38.5-40.8) | neg (no ct/27.8-30.5) | New Amplidiag® software version interprets the result as negative even though there is amplification from orf1ab gene |
| 2 | neg (no ct/no ct) | pos (27.5-28.0/no ct-27.5) | Specimens positive by cobas® test |
| 2 | pos (no ct/38.0-39.6) | neg (no ct/no ct) |  |

### Performance of the new software version of Amplidiag COVID-19 test

Of the 147 specimens run with both the new and old software version of the Amplidiag® COVID-19 test 11 specimens gave failed result with version 1 or version 2 of the software or both (n=2) and were excluded from the analysis. 73 positive and 63 negative results were obtained by the old software and 68 positive and 68 negative results by the new version. There were 9 discrepant results. The specimen that gave false positive result by the earlier version of the Amplidiag software was negative in the new run with the new Amplidiag software. Four specimens that were positive with the old version gave negative result by the new version, even though there was amplification from the orf1ab target. An additional two specimens were positive with the old version and negative with the new version without amplification from orf1ab target. Two specimens negative by the old version, a positive result was obtained when rerun with the new software version. These two specimens were positive by cobas® assay only in the comparison of the three methods (Table 6B)

## Discussion

In general, all tested assays performed adequately. Both the time from sample to result and hands-on time per sample were shortest for cobas® test. The cobas® and Amplidiag® assays allow for an automatic transfer of the results to the laboratory information system. On the other hand, both LDT and Amplidag® allow for the evaluation of the actual amplification curves, which is very important for the quality assurance of the results, especially when the tests have been set up in a straining timetable.

Of the three RT-PCR methods evaluated in this study, cobas® SARS-CoV-2 test showed the best overall performance for the detection of SARS-CoV-2 RNA in clinical respiratory specimens. In the absence of a gold standard, the reference value was created as a consensus of two methods. The agreement between the different methods tested was excellent. However, the positive percent agreement was highest for cobas® SARS-CoV-2 test (100%), while its negative percent agreement was the lowest (89.4%). All the specimens positive only with cobas® SARS-Cov-2 test had high ct-values (>30), with one exception, where ct-values of 28.6 (orf1) and 29.1 (E-gene) were obtained. The dilution series conducted for specimens collected in Copan UTM vs. 0.9% saline, however, suggests significantly higher sensitivity for the cobas® assay in comparison with the other two assays and the low NPA value may well be due to the same reason. Previous studies also suggest high sensitivity for the cobas® SARS-CoV-2 test^8,9^. The second commercial test evaluated, the Amplidiag® COVID-19 test obtained same PPA (98.9%) as LDT, but lower NPA (98.8% vs. 100% respectively). There was one specimen that was positive by Amplidiag® COVID-19 test only, which seemed to be a false positive by interpretation of the amplification curve of the positive target (orf1ab). This specimen was negative by the new Amplidiag® software version, which did not give any false interpretation of negative results in this study. However, it seems that what is gained in specificity is lost in sensitivity: With the new software version three specimens positive by the old version gave negative results, even though specific amplification from the orf1ab target could be seen. All things considered, Amplidiag® test seems to perform on the same par with LDT, apart from the relatively high failure rate with Amplidiag® test (5.5%). The comparison of results from eNAT tubes versus mNAT tubes suggests that the Amplidiag® test is for some reason less sensitive (later ct value) for the sampling control (RNase P gene) from mNAT tubes. The difference in obtained ct values was greater for RNase P gene (∼2 ct cycles) than for the specific virus targets (<1 ct cycle). This was also observed in the rising failure percent (from 3.8 % to 8.4%, data not shown) when the change to mNAT tubes was implemented.

We analyzed the QCMD 2020 Coronavirus Outbreak Preparedness EQA Pilot Study with all the evaluated methods available in our laboratory. The cobas® test and LDT identified all samples correctly. Amplidiag® missed repeatedly one specimen with dPCR reference value of 3.3 log10 copies/ml, although it correctly identified another sample with dPCR reference value of 2.3 log10 copies/ml. In addition, Amplidiag® test yielded a failed test result for 4 samples, as these samples lacked human RNase P target needed for a valid negative result with the test. The lack of correct matrix in EQA samples is a major drawback in general. The performance of the test identifying the target virus may be very different in the correct matrix in comparison to virus alone. Human nucleic acids, if not yielding a false positive result in a PCR, may take force from the amplification reaction (e.g. if primers and/or probe have binding sites in human genome or transcriptome). For example, the LDT amplifies virus positive specimens without human matrix more efficiently than specimens with human matrix included (data not shown).

It is common that RNA-viruses accumulate mutations at high frequency since RNA-polymerase lacks proofreading activity. Indeed, there is evidence that also SARS-CoV-2 is evolving during time and mutations may occur in the target area of molecular tests used. A previous report suggests that a mutation has already occurred in the target region of the cobas® SARS-CoV-2 E-gene test^10^. In this study, all specimens gave a positive signal for both target genes by cobas® test. Fortunately, the test uses a dual target approach, which means that the mutation proposed to be in the target area of the cobas® E-gene test does not compromise correct results. This highlights the importance of a dual target approach^11-13^.

Many countries are now (in July 2020) moving from lock-down to “test, trace and isolate” strategy in their COVID-19 mitigation, which requires a large capacity for sensitive and reliable SARS-CoV-2 laboratory testing. Clinical laboratories need to be prepared for uninterrupted high-throughput testing during the coming months. In order to achieve this, laboratories need to deploy optional testing methods for the event of breakage in instrumentation, as well as prepare for the ongoing global shortage in testing supplies. Therefore, it is of critical importance for clinical laboratories to maintain several simultaneous platforms in their SARS-CoV-2 nucleic acid testing, and continuously monitor the performance of the assays used.

## Data Availability

All data referred to in the manuscript is available

## Acknowledgments

The laboratory staff doing the SARS-CoV-2 diagnostics at HUSLAB are thanked for the extra work they have done for this evaluation during the straining times.

## REFERENCES

1. Cheng MP, Papenburg J, Desjardins M, Kanjilal S, Quach C, Libman M, Dittrich S, Yansouni CP. Diagnostic Testing for Severe Acute Respiratory Syndrome-Related Coronavirus-2: A Narrative Review. Ann Intern Med 2020, 13:M20–1301.

2. Jääskeläinen AJ, Kekäläinen E, Kallio-Kokko H, Mannonen L, Kortela E, Vapalahti O, Kurkela S, Lappalainen M. Evaluation of commercial and automated SARS-CoV-2 IgG and IgA ELISAs using coronavirus disease (COVID-19) patient samples. Euro Surveill 2020, 25:2000603.

3. Corman VM, Landt O, Kaiser M, Molenkamp R, Meijer A, Chu DK, Bleicker T, Brünink S, Schneider J, Schmidt ML, Mulders DG, Haagmans BL, van der Veer B, van den Brink S, Wijsman L, Goderski G, Romette JL, Ellis J, Zambon M, Peiris M, Goossens H, Reusken C, Koopmans MP, Drosten C. Detection of 2019 novel coronavirus (2019-nCoV) by real-time RT-PCR. Euro Surveill 2020, 25:2000045.

4. Kalorama Information, COVID-19 IVD Test Tracker: https://kaloramainformation.com/covid19diagnosticstracker/

5. Rodino KG, Espy MJ, Buckwalter SP, Walchak RC, Germer JJ, Fernholz E, Boerger A, Schuetz AN, Yao JD, Binnicker MJ. Evaluation of Saline, Phosphate-Buffered Saline, and Minimum Essential Medium as Potential Alternatives to Viral Transport Media for SARS-CoV-2 Testing. J Clin Microbiol 2020, 58:e00590–20.

6. Nummi M, Mannonen L, Puolakkainen M. Development of a multiplex real-time PCR assay for detection of Mycoplasma pneumoniae, Chlamydia pneumoniae and mutations associated with macrolide resistance in Mycoplasma pneumoniae from respiratory clinical specimens. Springerplus 2015, 4:684.

7. Konrad R, Eberle U, Dangel A, Treis B, Berger A, Bengs K, Fingerle V, Liebl B, Ackermann N, Sing A. Rapid establishment of laboratory diagnostics for the novel coronavirus SARS-CoV-2 in Bavaria, Germany, February 2020. Euro Surveill 2020, 25:2000173.

8. Poljak M, Korva M, Knap Gašper N, Fujs Komloš K, Sagadin M, Uršič T, Avšič Županc T, Petrovec M. Clinical evaluation of the cobas SARS-CoV-2 test and a diagnostic platform switch during 48 hours in the midst of the COVID-19 pandemic. J Clin Microbiol 2020, 58:e00599–20.

9. Pujadas E, Ibeh N, Hernandez MM, Waluszko A, Sidorenko T, Flores V, Shiffrin B, Chiu N, Young-Francois A, Nowak MD, Paniz-Mondolfi AE, Sordillo EM, Cordon-Cardo C, Houldsworth J, Gitman MR. Comparison of SARS-CoV-2 detection from nasopharyngeal swab samples by the Roche cobas 6800 SARS-CoV-2 test and a laboratory-developed real-time RT-PCR test. J Med Virol 2020, May 8. doi: 10.1002/jmv.25988. Online ahead of print.

10. Artesi M, Bontems S, Göbbels P, Franckh M, Boreux R, Meex C, Melin P, Hayette M-P, Bours V, Durkin K. Failure of the cobas® SARS-CoV-2 (Roche) E-gene assay is associated with a C-to-T transition at position 26340 of the SARS-CoV-2 genome medRxiv preprint doi: https://doi.org/10.1101/2020.04.28.20083337.

11. Hokynar K, Rantakokko-Jalava K, Hakanen A, Havana M, Mannonen L, Jokela P, Kurkela S, Lappalainen M, Unemo M, Puolakkainen M. The Finnish New Variant of Chlamydia trachomatis with a Single Nucleotide Polymorphism in the 23S rRNA Target Escapes Detection by the Aptima Combo 2 Test. Microorganism. 2019, 7:227.

12. Mannonen L, Loginov R, Helanterä I, Dumoulin A, Vilchez RA, Cobb B, Hirsch HH, Lautenschlager I. Comparison of two quantitative real-time CMV-PCR tests calibrated against the 1st WHO international standard for viral load monitoring of renal transplant patients. J Med Virol 2014, 86:576–584.

13. Beckmann C, Dumoilin A, Rinaldo CH, Hirsch HH. Comparison of a UL111a realtime PCR and pp65 antigenemia for the detection of cytomegalovirus. J Med Virol 2011, 83:2143–2150.

